# Racial and Ethnic Disparities in Health of Adults in the United States: A 20-Year National Health Interview Survey Analysis, 1999-2018

**DOI:** 10.1101/2020.10.30.20223487

**Authors:** Shiwani Mahajan, César Caraballo, Yuan Lu, Dorothy Massey, Karthik Murugiah, Amarnath R. Annapureddy, Brita Roy, Carley Riley, Oyere Onuma, Marcella Nunez-Smith, Javier Valero-Elizondo, Howard P. Forman, Khurram Nasir, Jeph Herrin, Harlan M. Krumholz

**Affiliations:** Center for Outcomes Research and Evaluation, Yale New Haven Hospital, New Haven, CT; Section of Cardiovascular Medicine, Department of Internal Medicine, Yale School of Medicine, New Haven, CT; Section of General Internal Medicine, Department of Internal Medicine, Yale School of Medicine, New Haven, CT; Department of Chronic Disease Epidemiology, Yale School of Public Health, New Haven, CT; Department of Pediatrics, University of Cincinnati College of Medicine, Cincinnati, OH; Division of Critical Care Medicine, Cincinnati Children’s Hospital Medical Center, Cincinnati, OH; Equity Research and Innovation Center, General Internal Medicine, Yale School of Medicine, New Haven, CT; Division of Cardiovascular Prevention and Wellness, Houston Methodist DeBakey Heart and Vascular Center, Houston, TX; Center for Outcomes Research, Houston Methodist, TX; Department of Radiology and Biomedical Imaging, Yale School of Medicine, New Haven, CT; Department of Health Policy and Management, Yale School of Public Health, New Haven, CT

## Abstract

**Importance:** Thirty-five years ago, the Heckler Report described health disparities among minority populations in the US. Since then, policies have been implemented to address these disparities. However, a recent evaluation of progress towards improving the health and health equity among US adults is lacking.

**Objectives:** To evaluate racial/ethnic disparities in the physical and mental health of US adults over the last 2 decades.

**Design:** Cross-sectional.

**Setting:** National Health Interview Survey data, years 1999–2018.

**Participants:** Adults aged 18–85 years.

**Exposure:** Race/ethnicity subgroups (non-Hispanic White, non-Hispanic Black, non-Hispanic Asian, Hispanic).

**Main outcome and measures:** Proportion of adults reporting poor/fair health status, severe psychological distress, functional limitation, and insufficient sleep. We also estimated the gap between non-Hispanic White and the other subgroups for these four outcomes.

**Results:** We included 596,355 adults (mean age 46 years, 51.8% women), of which 69.7%, 13.8%, 11.8% and 4.7% identified as non-Hispanic White, Hispanic, non-Hispanic Black, and non-Hispanic Asian, respectively. Between 1999 and 2018, Black individuals fared worse on most measures of health, with 18.7% (95% CI 17.1–20.4) and 41.1% (95% CI 38.7–43.5) reporting poor/fair health and insufficient sleep in 2018 compared with 11.1% (95% CI 10.5– 11.7) and 31.2% (95% CI 30.3–32.1) among White individuals. Notably, between 1999–2018, there was no significant decrease in the gap in poor/fair health status between White individuals and Black (−0.07% per year, 95% CI −0.16–0.01) and Hispanic (−0.03% per year, 95% CI −0.07– 0.02) individuals, and an increase in the gap in sleep between White individuals and Black (+0.2% per year, 95% CI 0.1–0.4) and Hispanic (+0.3% per year, 95% CI 0.1–0.4) individuals. Additionally, there was no significant decrease in adults reporting poor/fair health status and an increase in adults reporting severe psychological distress, functional limitation, and insufficient sleep.

**Conclusions and Relevance:** The marked racial/ethnic disparities in health of US adults have not improved over the last 20 years. Moreover, the self-perceived health of US adults worsened during this time. These findings highlight the need to re-examine the initiatives seeking to promote health equity and improve health.

## INTRODUCTION

The landmark 1985 Report of the United States (US) Department of Health and Human Services’ (HHS) Task Force on Black and Minority Health released in 1985, commonly known as the Heckler Report, described health disparities and sought to stimulate efforts to eliminate them.^1^ Since then, the US HHS established an Office of Minority Health to introduce programs and policies to address health disparities.^2^ The Centers for Disease Control and Prevention and the Agency for Healthcare Research and Quality also prioritized activities to reduce health disparities.^3,4^ Similarly, national programs, such as Healthy People 2000, 2010, and 2020 set goals to eliminate disparities in health outcomes.^5^ Now, 35 years since the publication of the Heckler Report, is an opportune time to assess what the nation has achieved through these programs in bridging these gaps.

Previous studies have reported on physical and mental health indicators across racial and ethnic subgroups in the US.^6-14^ However, most reports have used older or shorter study periods or have not had representative samples. Additionally, prior studies provide varied estimates on the trends in these health indicators based on the years evaluated. As such, we lack a recent comprehensive review of how the physical and mental health status of different racial and ethnic subgroups has changed over the last two decades.

Accordingly, we leveraged data from the National Health Interview Survey (NHIS),^15^ which provides annual estimates on a broad range of health measures based on a large sample of the population in the US, to evaluate our progress towards improving health and health equity between 1999 and 2018. Specifically, we evaluated trends in self-perceived general health status, psychological stress, physical function, and sleep sufficiency as health indicators across racial and ethnic subgroups.

## METHODS

### Study design and population

We included data from 603,140 adults, aged 18–85 years, included in the NHIS for years 1999–2018. The NHIS is an annual, cross-sectional, weighted, multi-stage sampled survey that provides nationally-representative estimates on the non-institutionalized US population.^15^ Individuals were categorized into four mutually exclusive subgroups based on their self-reported race/ethnicity information: non-Hispanic White, non-Hispanic Black, non-Hispanic Asian and Hispanic subgroups. We excluded individuals with missing information on race/ethnicity (n=112) and those who identified as Alaskan Native or American Indian or ‘Other’ Race (n=6,673) due to small numbers (**eFigure1**). For simplicity, we hereafter refer to these groups as White, Black, Asian, and Hispanic.

We obtained the NHIS data from the Integrated Public Use Microdata Series Health Surveys that compiles the annual NHIS microdata, simplifying access to data across multiple years and facilitating consistent comparisons across time periods.^16^ This study received an exemption for review from the Institutional Review Board at Yale University because NHIS data are publicly available and deidentified. The study was reported following the Strengthening the Reporting of Observational Studies in Epidemiology reporting guidelines.

### Outcome variables

#### Health status

Data on health status were available for all years. Health status was assessed using the question: “Would you say your health in general is excellent, very good, good, fair, or poor?” Responses were rated on a five-point Likert scale, ranging from “excellent” to “poor.” Self-assessed health status has been validated as a useful indicator of health for a variety of populations and allows for broad comparisons across different conditions and populations.^17,18^

#### Mental health

Data on mental health were available for all years. Mental health was assessed using the Kessler-6 Scale (K6), which asks about 6 manifestations of nonspecific psychological distress over a 30-day recall period.^19^ The K6 asks how often, during the past 30 days, the respondent felt: 1) so sad that nothing could cheer him/her up, 2) nervous, 3) restless or fidgety, 4) hopeless, 5) that everything was an effort, and 6) worthless. Reponses were rated on a 5-point scale based on the reported frequency of the feelings and were each assigned 0 to 4 points (i.e., 0 for “none of the time”; 1 for “a little of the time”; 2 for “some of the time”; 3 for “most of the time”; and 4 for “all of the time”). The summed responses ranged from 0 to 24 and persons with a score of ≥13 were identified as having severe psychological distress.

#### Physical functioning

Data on physical functioning were available for all years. Physical functioning was assessed by asking the participants how difficult it was for them, by themselves and without using any special equipment, to: 1) walk a quarter of a mile (or about 3 city blocks), 2) walk up 10 steps without resting, 3) stand or be on their feet for about 2 hours, 4) sit for about 2 hours, 5) stoop, bend, or kneel, 6) reach up over their head, 7) use their fingers to grasp or handle small objects, 8) lift or carry something as heavy as 10 pounds (such as a full bag of groceries), 9) push or pull large objects like a chair, 10) go out to things like shopping, movies, or sporting events, 11) participate in social activities such as visiting friends, attending clubs and meetings, going to parties, and 12) do things to relax at home or for leisure (such as reading, watching tv, sewing). Responses for each of these 12 questions were rated on a 6-point scale: not at all difficult, only a little difficult, somewhat difficult, very difficult, can’t do at all, do not do this activity. We assessed the proportion of adults who were “limited in any way” by identifying participants that reported any degree of difficulty in any one or more of the functional activities.^7^

#### Sleep duration

Information on hours of sleep was available for years 2004 to 2018. Hours of sleep were assessed by asking the participants: “On average, how many hours of sleep do you get in a 24-hour period?” Hours of sleep were recorded in whole numbers, rounding ≥30 minutes up to the next hour. We defined insufficient sleep as <7 hours of sleep in a 24-hour period based on the recommendations of the American Academy of Sleep Medicine and Sleep Research Society.^20^

### Other sociodemographic variables

Age (in years) was included as a covariate. Other variables described include: sex (male, female), education level (less than high school, high school diploma or GED, some college, Bachelor’s degree or higher), family income (based on percent of family income relative to the federal poverty limit from the Census Bureau: high/middle income [≥200%] and low-income [<200%]), insurance status (insured, uninsured), geographic region (Northeast, Midwest, South, West), citizenship status (US citizen, non-US citizen), marital status (married, unmarried), employment status (working, not in labor force, unemployed), comorbidities (hypertension, diabetes, prior stroke or myocardial infarction, cancer, emphysema or chronic bronchitis), smoking status, obesity, and flu vaccination in the past 12 months. Information on all sociodemographic variables was available for all years and responses coded as “unknown” or “not ascertained” were analyzed under a separate category of “unknown.”

### Statistical analysis

First, we summarized respondent characteristics by race/ethnicity for the first two (1999 and 2000) and last two (2017 and 2018) years of the study, separately. Then, to examine trends, we first estimated the annual age-adjusted, survey-weighted proportion of adults with poor/fair health status, severe psychological distress, functional limitation, and insufficient sleep for each subgroup of race/ethnicity using a linear probability model, with the outcome as the dependent variable, age (discrete) as independent variables, and an indicator for each year of interview. Age was centered on its overall mean for the study sample and the intercept was suppressed; the coefficients for each year then represented age-adjusted annual outcome rates. We then estimated trends in outcomes by fitting regression models for each race/ethnicity subgroup, with an age-adjusted annual prevalence of outcome being the dependent variable and time in years as the independent variable. To account for the serial correlation of annual outcome rates, we incorporated an autoregressive error term, with 1-year correlation, and used the coefficient of the time variable to evaluate the slope of each outcome’s prevalence during the study period. Next, we estimated the absolute prevalence difference of each outcome by race/ethnicity between 1999 and 2018.

Finally, we assessed the trends in the disparities across racial/ethnic subgroups by fitting regression models for Black, Asian, and Hispanic subgroups to assess if the gap with the White subgroup had changed significantly between 1999 and 2018. All analyses accounted for the complex survey design of the NHIS, including appropriate sampling weights and sampling units, to ensure that our results were nationally representative. We considered 2-sided *P*-values <0.05 to be statistically significant. All analyses were performed using Stata SE version 16.0 (StataCorp, College Station, TX).

## RESULTS

### Population characteristics

Our final study sample included 596,355 adults, with mean (standard error) age of 46.2 (0.1) years and 51.8% were women. Of these, 69.7% adults identified as White individuals (mean age 48.1 [0.1] years; 51.7% women), 11.8% as Black individuals (mean age 43.4 [0.1] years; 55.2% women), 4.7% as Asian individuals (mean age 43.6 [0.2] years; 52.3% women), and 13.8% as Hispanic individuals (40.3 [0.1] years; 49.6% women). The study population’s sociodemographic and clinical characteristics by race/ethnicity during the first two years (1999 and 2000) and last two years (2017 and 2018) of the study period are shown in **eTable 1**.

**Table 1.** Trends in poor/fair health status, severe psychological distress, insufficient sleep, and functional limitation by race/ethnicity.

| **Outcome** | **Non-Hispanic White,**<br>**% (95% CI)** | **Hispanic,**<br>**% (95% CI)** | **Non-Hispanic Black,**<br>**% (95% CI)** | **Non-Hispanic Asian,**<br>**% (95% CI)** |
| --- | --- | --- | --- | --- |
| **Poor/Fair health status** |  |  |  |  |
| Absolute difference in prevalence 1999-2018 | 0.2 (-0.6 – 1.0) | 0.1 (-1.7 – 1.9) | -1.9 (-4.1 – 0.3) | -1.7 (-4.4 – 1.0) |
| Annualized rate of change in prevalence | -0.00 (-0.05 – 0.05) | -0.03 (-0.08 – 0.03) | -0.08 (-0.20 – 0.03) | -0.01 (-0.08 – 0.05) |
| Annualized rate of change in gap with non-Hispanic Whites | N/A | -0.03 (-0.07 – 0.02) | -0.07 (-0.16 – 0.01) | -0.01 (-0.10 – 0.07) |
| **Severe psychological distress** |  |  |  |  |
| Absolute difference in prevalence 1999-2018 | 1.5 (1.0 – 2.0) | 1.4 (0.4 – 2.4) | 1.3 (0.0 – 2.5) | 1.1 (-0.0 – 2.2) |
| Annualized rate of change in prevalence | 0.05 (0.03 – 0.07) | 0.03 (-0.00 – 0.07) | 0.03 (0.00 – 0.06) | 0.00 (-0.04 – 0.05) |
| Annualized rate of change in gap with non-Hispanic Whites | N/A | -0.02 (-0.07 – 0.03) | -0.02 (-0.05 – 0.01) | -0.05 (-0.1 – 0.01) |
| **Functional limitation** |  |  |  |  |
| Absolute difference in prevalence 1999-2018 | 7.7 (6.5 – 8.9) | 7.6 (5.3 – 10.0) | 7.4 (4.7 – 10.2) | 3.7 (-0.0 – 7.3) |
| Annualized rate of change in prevalence | 0.28 (0.16 – 0.41) | 0.34 (0.16 – 0.52) | 0.34 (0.21 – 0.48) | 0.15 (-0.04 – 0.33) |
| Annualized rate of change in gap with non-Hispanic Whites | N/A | 0.06 (-0.02 – 0.14) | 0.06 (-0.01 – 0.13) | -0.15 (-0.27 – -0.02) |
| **Insufficient sleep** |  |  |  |  |
| Absolute difference in prevalence 2004-2018 | 3.1 (2.0 – 4.3) | 6.3 (3.9 – 8.8) | 6.3 (3.3 – 9.4) | 1.4 (-3.0 – 5.8) |
| Annualized rate of change in prevalence | 0.27 (0.10 – 0.44) | 0.52 (0.30 – 0.74) | 0.53 (0.35 – 0.71) | 0.16 (-0.15 – 0.47) |
| Annualized rate of change in gap with non-Hispanic Whites | N/A | 0.27 (0.13 – 0.40) | 0.24 (0.07 – 0.41) | -0.11 (-0.31 – 0.09) |
| Abbreviations: CI, confidence interval |  |  |  |  |

### Trends and disparities in health status

Overall, 595,952 (99.9%) individuals had complete information on their health status. Between 1999 and 2018, Black and Hispanic individuals had a significantly higher age-adjusted proportion of adults reporting poor/fair health compared with White and Asian individuals, with 18.7% (95% CI 17.1%–20.4%), 17.8% (95% CI 16.4%–19.2%), 11.1% (95% CI 10.5%–11.7%), and 11.0% (95% CI 9.2%–12.8%) of Black, Hispanic, White, and Asian individuals, respectively, reporting poor/fair health status in 2018 **(Figure 1)**. Notably, between 1999 and 2018, there was no significant decrease in the gap in poor/fair health status between White individuals and Black (−0.07% per year, 95% CI −0.16%–0.01%) and Hispanic (−0.03% per year, 95% CI −0.07%–0.02%) individuals. Additionally, there was no significant difference in the prevalence of adults with poor/fair health status between 1999 and 2018 among White (+0.2%, 95% CI −0.6%–1.0%), Black (−1.9%, 95% CI −4.1%–0.3%), Hispanic (+0.1%, 95% CI −1.7%– 1.9%), or Asian (−1.7%, 95% CI −4.4%–1.0%) subgroups (**Table 1**). The annualized rate of change in prevalence of adults with poor/fair health status was also not significant for any of the four subgroups.

**Figure 1.**
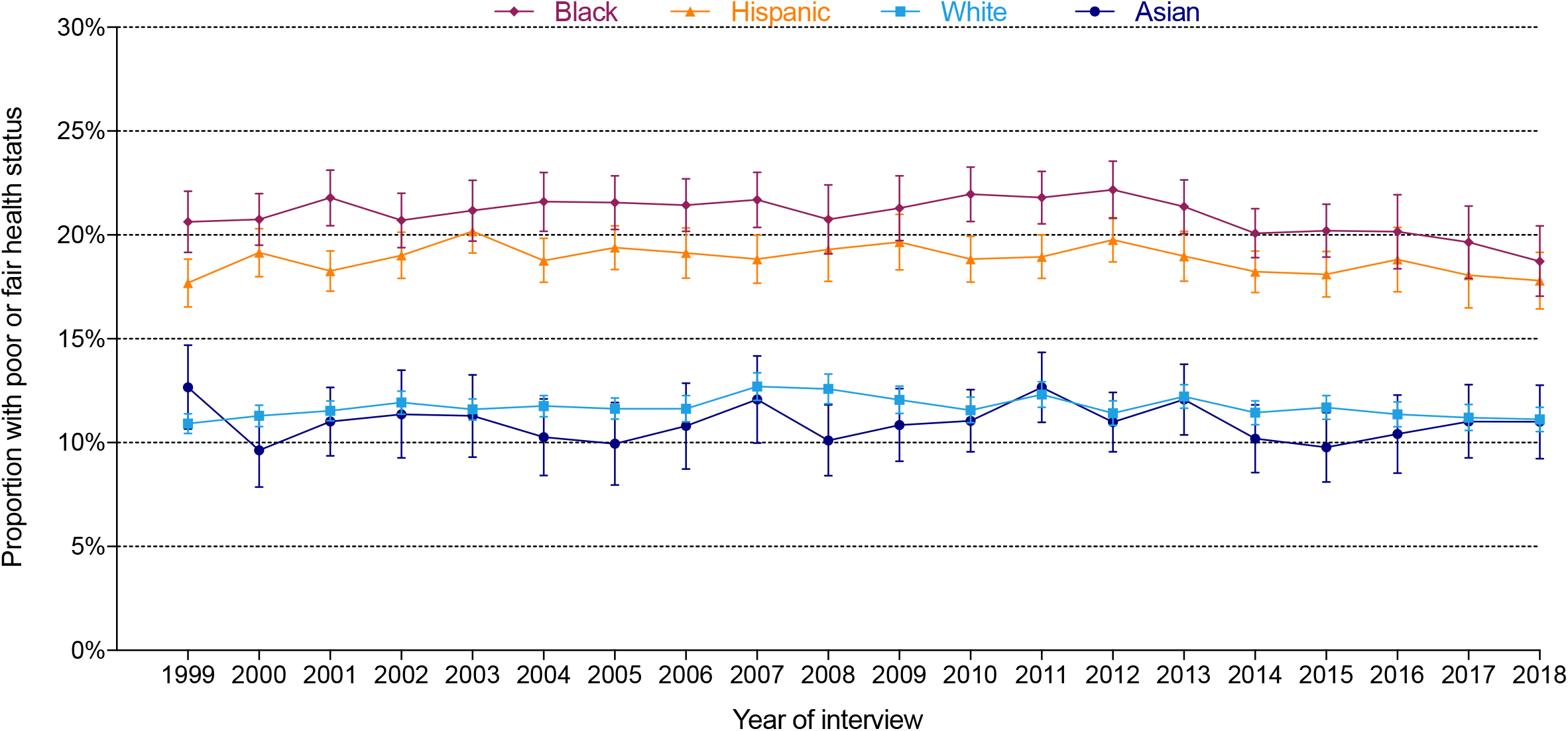
Annual age-adjusted proportion of individuals reporting poor/fair health status among adults in the US, by race/ethnicity.

### Trends and disparities in mental health

Overall, 582,892 (97.7%) individuals provided complete information on their mental health. Between 1999 and 2018, there was no significant difference in the age-adjusted proportion of individuals with severe psychological distress by race/ethnicity (**Figure 2**). The absolute prevalence of adults reporting severe psychological distress increased between 1999 and 2018 among White (+1.5%, 95% CI 1.0%–2.0%), Black (+1.3%, 95% CI 0.0%–2.5%), and Hispanic (+1.4%, 95% CI 0.4%–2.4%) individuals, but remained stable for Asian (+1.1%, 95% CI −0.0%–2.2%) subgroup (**Table 1**). The annualized rate of change in prevalence of severe psychological distress increased significantly among White (+0.05% per year, 95% CI 0.03%– 0.07%) and Black (+0.03% per year, 95% CI 0.00%–0.06%) individuals, remaining stable for the Hispanic (+0.03% per year, 95% CI −0.00%–0.07%) and Asian (+0.00% per year, 95% CI - 0.04%–0.05%) subgroups.

**Figure 2.**
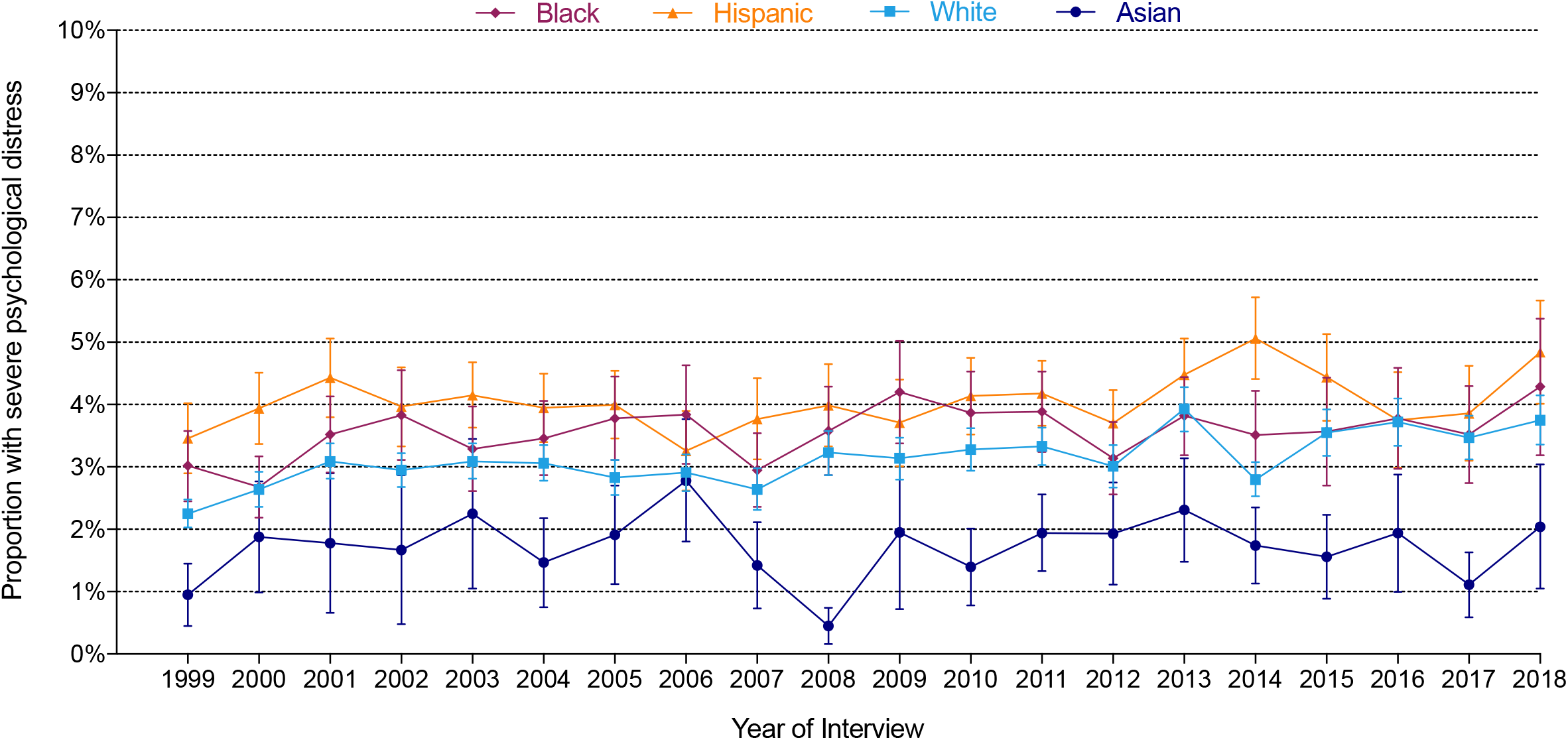
Annual age-adjusted proportion of individuals with severe psychological distress among adults in the US, by race/ethnicity.

### Trends and disparities in physical functioning

Overall, 594,691 (99.7%) individuals provided complete information on their physical functioning. Between 1999 and 2018, Black and White subgroups had the highest age-adjusted proportion of adults reporting functional limitation, followed by Hispanic and Asian subgroups, respectively (**Figure 3**). The absolute prevalence of functional limitation between 1999 and 2018 increased among White (+7.7%, 95% CI 6.5%–8.9%), Black (+7.4%, 95% CI 4.7%–10.2%), and Hispanic (+7.6%, 95% CI 5.3%–10.0%) individuals, remaining stable for Asian (+3.7%, 95% CI −0.0%–7.3%) subgroup (**Table 1**). The annualized change in adults reporting any functional limitation was +0.3% per year (95% CI 0.2%–0.4%) among White individuals, +0.3% per year (95% CI 0.2%–0.5%) among Black individuals, and +0.3% per year (95% CI 0.2%–0.5%) among Hispanic individuals.

**Figure 3.**
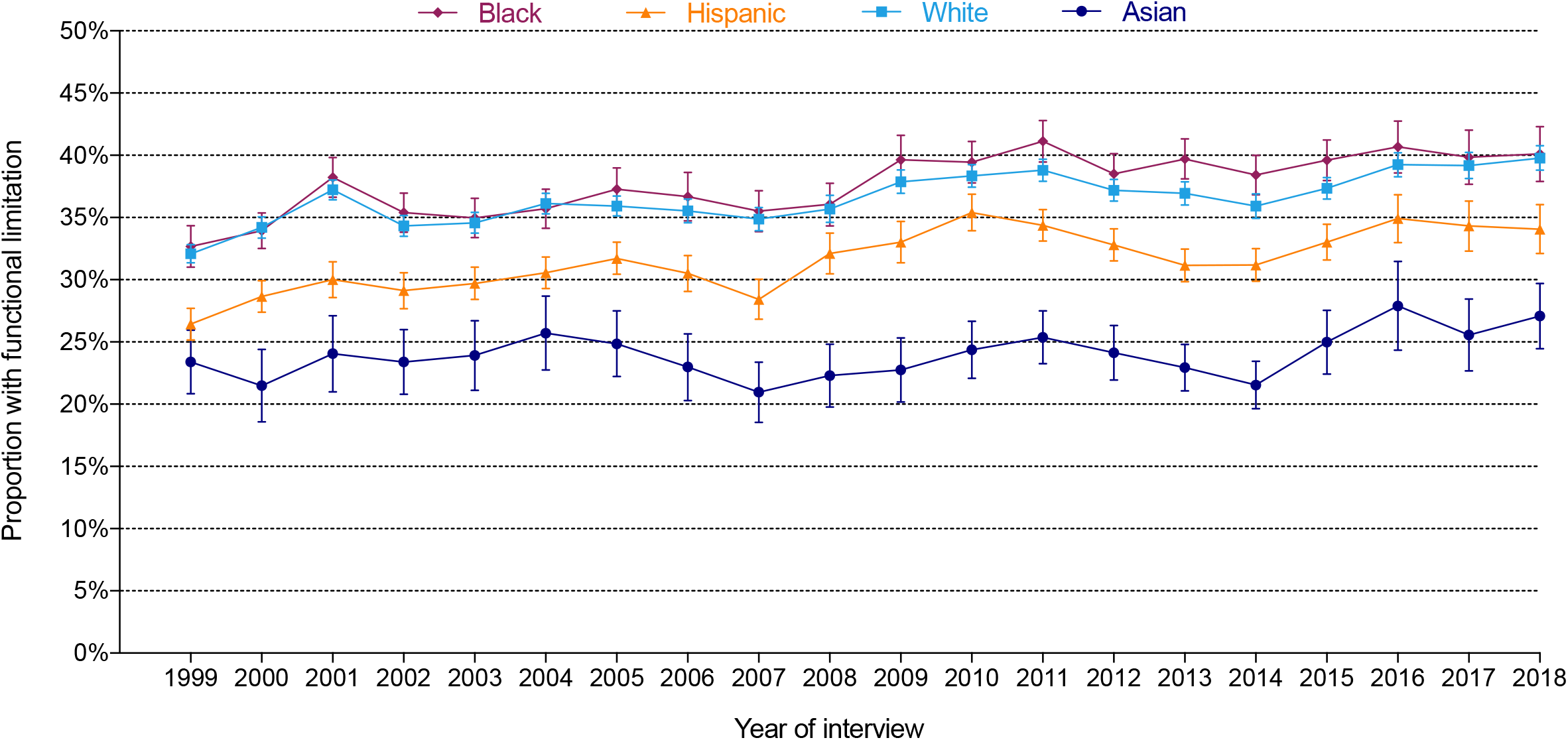
Annual age-adjusted proportion of individuals with functional limitation among adults in the US, by race/ethnicity.

### Trends and disparities in sleep sufficiency

Overall, 398,866 (97.7%) of the eligible individuals (n=408,347; interviewed between 2004–2018) provided complete information on their sleep hours. The overall distribution of hours of sleep by race/ethnicity is shown in **eFigure 2**. Between 2004 and 2018, Black and Asian individuals had significantly higher proportion of adults reporting insufficient sleep compared with White and Hispanic individuals **(Figure 4)**. Notably, the gap between White individuals and Black and Hispanic individuals increased by +0.2% per year (95% CI 0.1%–0.4%) and +0.3% per year (95% CI 0.1%–0.4%) during this time (**Table 1**). Additionally, the absolute prevalence of insufficient sleep during that period increased among White (+3.1%, 95% CI 2.0%–4.3%), Black (+6.3%, 95% CI 3.3%–9.4%), and Hispanic (+6.3%, 95% CI 3.9%–8.8%) individuals, but remained stable among the Asian (+1.4%, 95% CI −3.0%–5.8%) subgroup. The annualized change in adults reporting insufficient sleep was +0.3% per year (95% CI 0.1%–0.4%) among White individuals, +0.5% per year (95% CI 0.4%–0.7%) among Black individuals, and +0.5% per year (95% CI 0.3%–0.7%) among Hispanic individuals.

**Figure 4.**
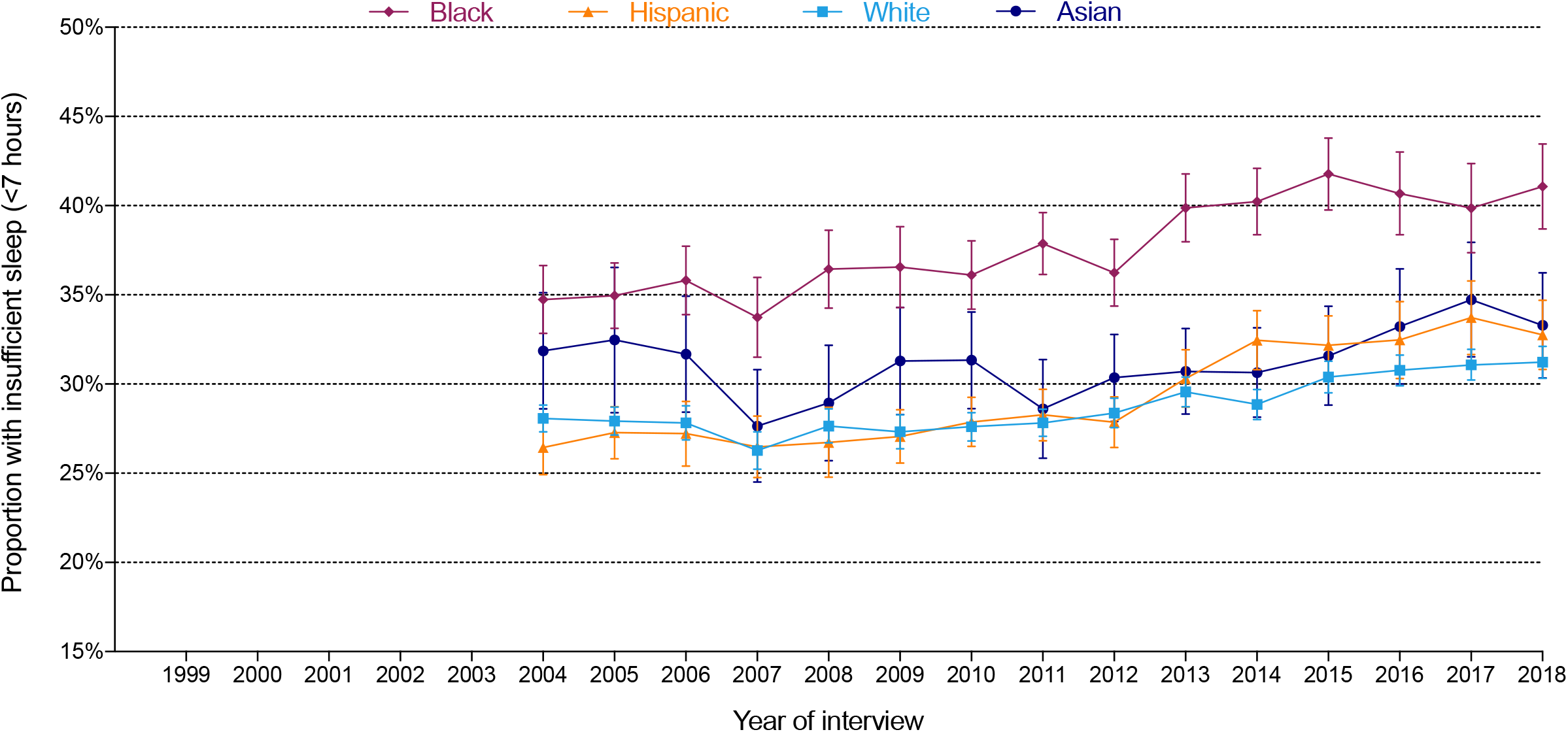
Annual age-adjusted proportion of individuals with insufficient sleep among adults in the US, by race/ethnicity.

## DISCUSSION

In this large, nationally representative study we report no progress in reducing disparities in health outcomes in the US. Moreover, the overall self-perceived health and well-being of adults living in the US have not improved in the past 20 years and may have actually worsened. We also report no decrease in adults reporting poor health status, and an increase in those reporting severe psychological distress, insufficient sleep, and functional limitation. This lack of change has occurred during a period of a massive increase in health care spending.^21^Our study expands the literature in several ways. We provide a comprehensive evaluation of trends in the self-perceived physical and mental health of different racial and ethnic groups in the US over the most recent 20 years. Though previous studies have described trends in health by race/ethnicity, most have focused on shorter time frames, which limits detailed evaluation of the progress we have made. For example, Zack et al.^22^ showed that between 1993 and 2001, though the overall percentage of adults with poor/fair health status increased by about 1.2% per year (95% CI 0.9% –1.6%), rates remained stable for White individuals, decreased slightly for Black individuals, and increased for Hispanic individuals. However, we found that over the last 2 decades there has been no significant decrease in the percentage of individuals reporting poor/fair health across any racial/ethnic subgroup. Similarly, studies looking at trends in severe psychological distress have shown conflicting results based on the years evaluated. For example, Mojtabai et al.^23^ found no significant trends in percentage of people reporting severe psychological distress between 2001 and 2012, whereas Olfson et al.^10^ reported a significant decline in severe psychological distress between 2004–2005 and 2014–2015. As such, our findings are important as they provide a long-term perspective of the trends over the last 20 years.

Our study also highlights the persistence of disparities in self-reported health between racial and ethnic subgroups in the US. We found that the gaps in these health measures have not only persisted but have worsened over the past two decades. These findings are consistent with previous reports that have shown a lack of progress on health equity in the US by race/ethnicity.^14^ Previous reports on the disparities in the health indicators assessed in this study also demonstrated no significant or little change in the race/ethnicity disparities in health status, psychological distress, short sleep duration, and physical functioning.^24-27^ Notably, consistent with prior reports, our findings show that Black individuals fare worse on most measures of health, which highlights that we have made little progress toward our goal of promoting health equity for minority groups, even 35 years after the release of the Heckler Report.

Our study has important public health and policy implications. Despite all the efforts to eliminate disparities in the overall health and well-being of the nation, we find no evidence of progress. As such, our findings highlight both the need for national accountability of our efforts towards promoting health equity, and for continued and more stringent efforts at the federal, regional, and local levels to reduce health disparities in the US. Additionally, policy changes to dismantle structural racism, and its inextricable downstream effects on physical and mental health, are needed to close the health equity gaps. Moreover, we find that for all groups there are worsening trends in the physical and mental health of the population. These findings suggest that, in addition to a failure to eliminate disparities, our current course is producing worse health outcomes, consistent with recent reports of a decline in life expectancy.^28^

Our study has several limitations. First, data in NHIS are self-reported and as such, may be subject to recall bias. However, self-reported data offer an accurate reflection of individuals’ perception of their own health. Second, response to self-perceived health status may be affected by an individual’s overall happiness. However, self-perceived health status has been validated as a reliable indicator of health across different populations. Third, though the K-6 correlates with several serious mental disorders, it includes only depression and anxiety-related symptoms, and several key dimensions of severe psychopathology are not represented. Fourth, NHIS does not distinguish between weekday and weekend sleep duration. Fifth, perceptions of healthful effects of adequate sleep may have influenced reporting of sleep duration due to social desirability bias. Finally, we did not analyze race groups with smaller representation such as Alaskan Native or American Indian because of inadequate sample size.

In conclusion, findings from this study indicate that marked racial and ethnic disparities in self-reported health have not improved over the last 20 years. Moreover, the self-perceived health of the US population has not only remained stagnant, but there is evidence that it has worsened in the last two decades, with no decrease in the percentage of the population reporting poor/fair health and an increased proportion reporting functional limitation, severe psychological distress, and insufficient sleep duration. These findings highlight the urgent need to re-examine the initiatives and policies that seek to eliminate disparities and improve health but that have thus far been inadequate.

## Data Availability

NHIS data are publicly available.

