## Supplemental Material for "Racial and Ethnic Disparities in Health of Adults in the United States: A 20-Year National Health Interview Survey Analysis, 1999-2018"

**Table of contents:**

eFigure 1. Flowchart of study population.

eFigure 2. Distribution of hours of sleep per day by race/ethnicity.

eTable 1. Sociodemographic and clinical characteristics of the study population during the first two years (1999 and 2000) and the last two years (2017 and 2018) of the study period by race/ethnicity.

**eFigure 1. Flowchart showing selection of the study population.**

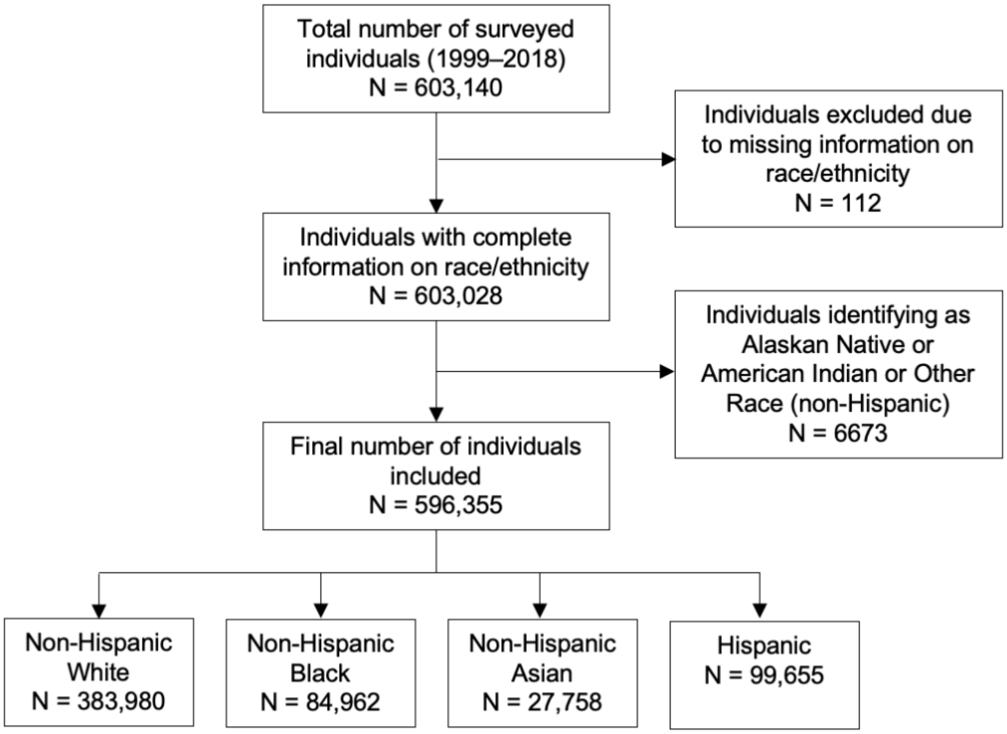

**eFigure 2. Distribution of hours of sleep per day by race/ethnicity.**

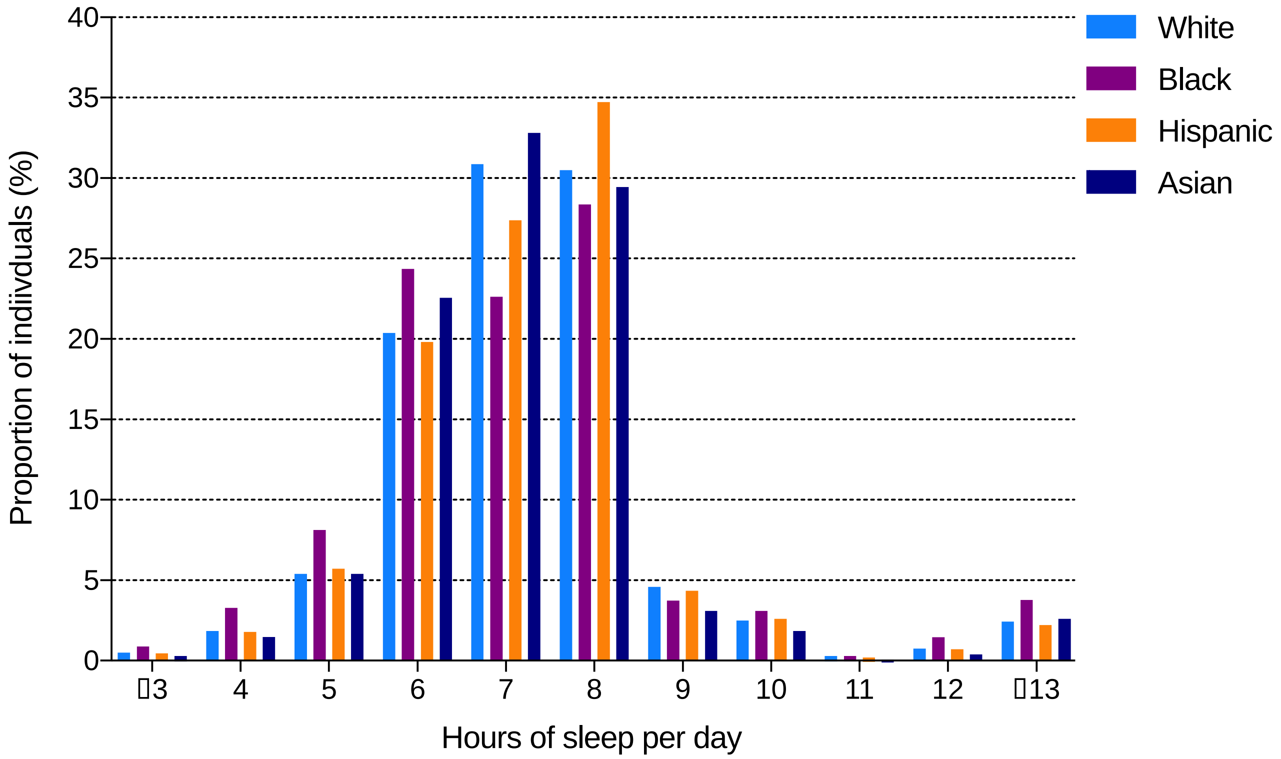

**eTable 1. Sociodemographic and clinical characteristics of the study population during the first two years (1999 and 2000) and the last two years (2017 and 2018) of the study period by race/ethnicity.**

| | Non-Hispanic White | | Hispanic | | Non-Hispanic Black | | Non-Hispanic Asian | |
|---|---|---|---|---|---|---|---|---|
| | 1999-2000 | 2017-2018 | 1999-2000 | 2017-2018 | 1999-2000 | 2017-2018 | 1999-2000 | 2017-2018 |
| **Characteristics** | n (%, 95% CI) | n (%, 95% CI) | n (%, 95% CI) | n (%, 95% CI) | n (%, 95% CI) | n (%, 95% CI) | n (%, 95% CI) | n (%, 95% CI) |
| **Total** | 41,882 | 36,410 | 10,305 | 6,423 | 8,758 | 5,884 | 1,597 | 2,640 |
| **Age (years)** | | | | | | | | |
| 18–39 | 15,169 (39.5% [38.9–40.2]) | 10,050 (33.3% [32.6–34.1]) | 3,610 (37.1% [35.0–39.2]) | 2,886 (50.1% [48.4–51.8]) | 3,862 (49.8% [48.3–51.3]) | 1,994 (43.7% [41.8–45.6]) | 844 (52.4% [49.2–55.6]) | 1,085 (41.9% [39.3–44.6]) |
| 40–64 | 17,246 (42.2% [41.5–42.8]) | 14,919 (42.3% [41.6–43.0]) | 6,617 (62.1% [60.0–64.2]) | 2,492 (39.1% [37.6–40.6]) | 3,539 (38.5% [37.1–39.8]) | 2,434 (41.1% [39.3–42.9]) | 581 (38.3% [35.1–41.4]) | 1,005 (42.5% [40.2–44.9]) |
| ≥65 | 9,467 (18.3% [17.8–18.8]) | 11,441 (24.4% [23.7–25.1]) | 78 (0.8% [0.6–1.1]) | 1,045 (10.8% [9.9–11.8]) | 1,357 (11.8% [11.0–12.6]) | 1,456 (15.3% [14.2–16.3]) | 172 (9.4% [7.8–11.4]) | 550 (15.5% [13.9–17.4]) |
| **Sex** | | | | | | | | |
| Men | 18,512 (48.1% [47.6–48.7]) | 16,798 (48.6% [48.0–49.3]) | 4,399 (49.4% [48.2–50.6]) | 2,788 (49.7% [48.1–51.4]) | 3,255 (44.4% [43.0–45.9]) | 2,435 (45.5% [43.7–47.2]) | 726 (48.7% [45.4–51.9]) | 1,270 (46.7% [44.4–49.0]) |
| Women | 23,370 (51.9% [51.3–52.4]) | 19,612 (51.4% [50.7–52.0]) | 5,906 (50.6% [49.4–51.8]) | 3,635 (50.3% [48.7–51.9]) | 5,503 (55.6% [54.1–57.0]) | 3,449 (54.5% [52.8–56.3]) | 871 (51.4% [48.1–54.6]) | 1,370 (53.3% [51.1–55.6]) |
| **Citizenship status** | | | | | | | | |
| US citizen | 41,177 (98.3% [98.2–98.5]) | 35,943 (98.4% [98.2–98.6]) | 6,617 (62.1% [60.0–64.2]) | 4,694 (71.5% [69.8–73.3]) | 8,385 (95.2% [94.5–95.8]) | 5,655 (94.8% [93.6–95.8]) | 921 (57.1% [53.5–60.6]) | 1,844 (69.9% [67.2–72.4]) |
| Non-US citizen | 672 (1.6% [1.5–1.8]) | 446 (1.5% [1.3–1.7]) | 3,610 (37.1% [35.0–39.2]) | 1,664 (27.4% [25.7–29.1]) | 360 (4.6% [4.0–5.3]) | 220 (5.1% [4.1–6.2]) | 658 (41.6% [38.0–45.2]) | 775 (29.2% [26.7–31.8]) |
| Unknown | 33 (0.0% [0.0–0.0]) | 21 (0.0% [0.0–0.1]) | 78 (0.8% [0.6–1.1]) | 65 (1.1% [0.8–1.6]) | 13 (0.2% [0.0–0.4]) | 9 (0.1% [0.0–0.3]) | 18 (1.3% [0.7–2.4]) | 21 (0.9% [0.5–1.7]) |
| **Education level** | | | | | | | | |
| Less than high school | 5,770 (13.3% [12.8–13.8]) | 2,708 (7.4% [6.9–7.9]) | 4,810 (44.0% [42.4–45.7]) | 1,831 (27.5% [25.7–29.4]) | 2,228 (24.1% [22.8–25.5]) | 948 (14.1% [12.8–15.6]) | 179 (11.2% [9.3–13.5]) | 221 (7.9% [6.6–9.4]) |

| | | | | | | | | |
|---|---|---|---|---|---|---|---|---|
| High school diploma /GED | 12,918 (31.5% [30.8–32.1]) | 8,710 (23.6% [22.8–24.3]) | 2,394 (24.0% [22.9–25.0]) | 1,615 (26.4% [24.8–28.0]) | 2,569 (31.1% [29.8–32.4]) | 1,646 (28.7% [27.1–30.3]) | 276 (17.7% [15.8–19.8]) | 383 (15.2% [13.4–17.3]) |
| Some college | 12,147 (29.3% [28.8–29.8]) | 11,509 (31.3% [30.5–32.0]) | 2,086 (21.4% [20.2–22.6]) | 1,737 (27.8% [26.3–29.4]) | 2,558 (29.3% [28.0–30.6]) | 1,905 (32.7% [31.0–34.5]) | 367 (23.8% [21.3–26.5]) | 546 (20.6% [18.7–22.8]) |
| ≥Bachelor's degree | 10,685 (25.2% [24.5–25.9]) | 13,386 (37.5% [36.4–38.6]) | 888 (9.2% [8.4–10.1]) | 1,193 (17.3% [15.9–18.8]) | 1,300 (14.3% [13.2–15.5]) | 1,347 (23.9% [22.0–25.9]) | 750 (45.6% [42.6–48.6]) | 1,478 (55.6% [52.5–58.6]) |
| Unknown | 362 (0.8% [0.7–0.9]) | 97 (0.3% [0.2–0.4]) | 127 (1.4% [1.1–1.8]) | 47 (1.0% [0.7–1.5]) | 103 (1.3% [1.0–1.6]) | 38 (0.6% [0.4–0.9]) | 25 (1.8% [1.2–2.7]) | 12 (0.7% [0.3–1.5]) |
| **Family income** | | | | | | | | |
| High/middle income | 30,643 (77.0% [76.4–77.6]) | 27,211 (78.8% [77.9–79.6]) | 4,412 (49.3% [47.6–50.9]) | 3,165 (53.5% [51.4–55.5]) | 4,372 (54.5% [52.5–56.5]) | 2,961 (55.3% [52.9–57.6]) | 1,082 (71.8% [68.3–75.1]) | 1,886 (73.8% [71.0–76.5]) |
| Low income | 11,239 (23.0% [22.4–23.6]) | 9,199 (21.2% [20.4–22.1]) | 5,893 (50.7% [49.1–52.4]) | 3,258 (46.5% [44.5–48.6]) | 4,386 (45.5% [43.6–47.5]) | 2,923 (44.7% [42.4–47.1]) | 515 (28.2% [24.9–31.7]) | 754 (26.2% [23.5–29.0]) |
| **Insurance status** | | | | | | | | |
| Insured | 37,216 (88.7% [88.3–89.1]) | 34,050 (93.2% [92.8–93.6]) | 6,669 (63.4% [61.6–65.2]) | 4,937 (75.4% [73.4–77.2]) | 7,036 (78.9% [77.8–80.0]) | 5,216 (87.0% [85.6–88.3]) | 1,329 (82.3% [80.0–84.5]) | 2,469 (93.2% [91.8–94.3]) |
| Uninsured | 4,522 (11.0% [10.6–11.4]) | 2,270 (6.5% [6.2–6.9]) | 3,591 (36.1% [34.3–37.9]) | 1,454 (23.8% [22.0–25.8]) | 1,663 (20.1% [19.1–21.2]) | 632 (11.9% [10.7–13.4]) | 260 (17.3% [15.1–19.7]) | 163 (6.3% [5.2–7.6]) |
| Unknown | 144 (0.3% [0.3–0.4]) | 90 (0.3% [0.2–0.4]) | 45 (0.5% [0.4–0.7]) | 32 (0.8% [0.5–1.3]) | 59 (1.0% [0.7–1.3]) | 36 (1.1% [0.7–1.7]) | 8 (0.4% [0.2–0.9]) | 8 (0.6% [0.3–1.2]) |
| **Region** | | | | | | | | |
| Northeast | 8,168 (20.2% [19.5–21.0]) | 6,431 (19.2% [17.5–21.1]) | 1,639 (15.7% [14.5–17.1]) | 789 (13.5% [11.2–16.2]) | 1,582 (18.0% [16.7–19.4]) | 733 (16.0% [13.5–18.8]) | 333 (21.2% [18.7–24.0]) | 493 (20.8% [16.9–25.4]) |
| Midwest | 12,012 (29.4% [28.5–30.3]) | 10,273 (27.4% [25.7–29.3]) | 637 (7.9% [6.9–9.1]) | 640 (9.4% [7.6–11.6]) | 1,698 (18.8% [17.2–20.5]) | 914 (15.2% [13.0–17.8]) | 214 (14.7% [11.9–18.0]) | 325 (11.8% [9.5–14.6]) |
| South | 13,935 (33.9% [33.0–34.8]) | 11,884 (33.0% [30.9–35.2]) | 3,729 (35.4% [33.2–37.8]) | 2,614 (37.1% [32.5–42.0]) | 4,645 (55.7% [53.4–57.9]) | 3,754 (60.5% [56.7–64.3]) | 298 (18.9% [16.4–21.8]) | 683 (25.5% [21.6–30.0]) |
| West | 7,767 (16.5% [15.8–17.2]) | 7,822 (20.4% [18.3–22.7]) | 4,300 (40.9% [38.6–43.3]) | 2,380 (40.0% [35.3–44.8]) | 833 (7.6% [6.7–8.5]) | 483 (8.3% [6.9–9.9]) | 752 (45.2% [41.2–49.2]) | 1,139 (41.8% [36.7–47.1]) |
| **Marital status** | | | | | | | | |

| | | | | | | | | |
|---|---|---|---|---|---|---|---|---|
| Unmarried | 20,259 (38.3% [37.5–39.0]) | 19,152 (43.6% [42.8–44.3]) | 4,953 (41.3% [40.2–42.5]) | 3,569 (50.6% [48.9–52.2]) | 6,232 (62.2% [60.8–63.7]) | 4,364 (67.2% [65.5–68.9]) | 664 (35.5% [32.5–38.6]) | 1,123 (34.9% [32.5–37.4]) |
| Married/Living with partner | 21,431 (61.4% [60.6–62.1]) | 17,179 (56.3% [55.5–57.0]) | 5,302 (58.3% [57.1–59.4]) | 2,840 (49.1% [47.5–50.7]) | 2,459 (37.1% [35.7–38.6]) | 1,502 (32.6% [30.9–34.3]) | 923 (64.0% [61.0–67.0]) | 1,509 (64.9% [62.4–67.4]) |
| Unknown | 192 (0.3% [0.3–0.4]) | 79 (0.2% [0.1–0.2]) | 50 (0.4% [0.3–0.5]) | 14 (0.4% [0.2–0.6]) | 67 (0.6% [0.5–0.9]) | 18 (0.3 [0.2–0.4]) | 10 (0.5% [0.3–0.9]) | 8 (0.2% [0.0–0.5]) |
| **Employment Status** | | | | | | | | |
| With a job/Working | 26,251 (65.6% [65.0–66.3]) | 20,990 (62.1% [61.3–62.9]) | 6,430 (66.1% [64.9–67.4]) | 4,075 (66.5% [64.7–68.3]) | 5,410 (63.9% [62.4–65.4]) | 3,248 (61.0% [59.0–63.1]) | 1,029 (65.9% [63.2–68.5]) | 1,667 (67.1% [64.9–69.3]) |
| Not in labor force | 15,037 (32.9% [32.2–33.6]) | 14,581 (35.4% [34.6–36.2]) | 3,579 (31.1% [29.9–32.3]) | 2,138 (29.8% [28.0–31.8]) | 3,020 (31.9% [30.5–33.4]) | 2,308 (32.4% [30.5–34.3]) | 530 (31.7% [29.1–34.4]) | 907 (30.0% [27.8–32.3]) |
| Unemployed | 562 (1.4% [1.2–1.5]) | 824 (2.5% [2.3–2.7]) | 276 (2.5% [2.2–2.9]) | 208 (3.6% [3.0–4.3]) | 308 (3.9% [3.4–4.6]) | 324 (6.5% [5.7–7.4]) | 35 (2.2% [1.5–3.1]) | 64 (2.8% [2.1–3.7]) |
| Unknown | 32 (0.1% [0.1–0.1]) | 15 (0.0% [0.0–0.1]) | 20 (0.3% [0.2–0.4]) | 2 (0.02% [0.0–0.1]) | 20 (0.3% [0.2–0.4]) | 4 (0.1% [0.0–0.3]) | 3 (0.2% [0.0–0.8]) | 2 (0.1% [0.0–0.4]) |
| **Comorbidities** | | | | | | | | |
| Hypertension | 10,466 (23.0% [22.5–23.4]) | 13,280 (32.7% [31.9–33.4]) | 1,762 (15.3% [14.4–16.2]) | 1,673 (22.3% [21.0–23.8]) | 2,857 (28.8% [27.6–30.1]) | 2,687 (37.6 [35.9–39.3]) | 237 (14.9% [12.9–17.2]) | 670 (24.2% [22.2–26.3]) |
| Diabetes | 2,303 (5.2% [5.0–5.4]) | 3,698 (9.1% [8.7–9.4]) | 720 (6.3% [5.8–6.9]) | 766 (10.5% [9.6–11.4]) | 803 (8.2% [7.5–8.9]) | 888 (11.7% [10.8–12.7]) | 57 (3.9% [3.0–5.1]) | 247 (8.9% [7.7–10.3]) |
| Prior stroke/MI | 2,404 (5.1% [4.9–5.3]) | 2,762 (6.4% [6.1–6.8]) | 307 (2.5% [2.2–2.9]) | 266 (3.3% [2.8–3.9]) | 456 (4.5% [4.0–5.1]) | 484 (6.0% [5.4–6.7]) | 35 (1.7% [1.2–2.2]) | 97 (3.2% [2.5–4.0]) |
| Cancer | 3,628 (7.8% [7.5–8.1]) | 5,164 (12.3 [11.9–12.8]) | 235 (2.2% [1.9–2.5]) | 289 (3.4% [2.9–3.8]) | 301 (2.9% [2.5–3.4]) | 379 (4.8% [4.3–5.4]) | 31 (1.7% [1.1–2.7]) | 126 (4.3% [3.4–5.3]) |
| Emphysema / Chronic bronchitis | 2,785 (6.2% [5.9–6.5]) | 2,163 (5.3% [5.0–5.6]) | 330 (2.9% [2.5–3.4]) | 231 (2.8% [2.4–3.3]) | 444 (4.3% [3.8–4.7]) | 332 (4.4% [3.7–5.1]) | 35 (1.6% [1.1–2.4]) | 58 (2.0% [1.4–2.7]) |
| **Current smoker** | 10,085 (24.2% [23.7–24.8]) | 5,712 (15.1% [14.6–15.7]) | 1,915 (18.3% [17.4–19.3]) | 690 (9.8% [9.0–10.8]) | 2,124 (23.8% [22.6–25.1]) | 973 (14.7% [13.5–16.1]) | 247 (14.9% [12.9–17.2]) | 201 (7.2% [6.0–8.5]) |
| **Flu vaccine in past 12m** | 13,642 (30.7% [30.0–31.3]) | 17,708 (47.1% [46.4–47.9]) | 1,982 (18.6% [17.5–19.6]) | 2,379 (34.9% [33.3–36.5]) | 1,879 (20.6% [19.7–21.5]) | 2,198 (35.2% [33.4–37.1]) | 426 (26.1% [23.3–29.2]) | 1,270 (48.2% [45.6–50.7]) |

| | | | | | | | | |
|---|---|---|---|---|---|---|---|---|
| **Obese (BMI ≥30 kg/m²)** | 8,006 (20.2% [19.7–20.6]) | 10,627 (30.3% [29.6–31.0]) | 2,339 (23.0% [21.9–24.1]) | 2,093 (34.0% [32.5–35.6]) | 2,542 (29.1% [28.0–30.2]) | 2,312 (39.4% [37.8–41.1]) | 84 (6.1% [4.9–7.7]) | 289 (12.0% [10.5–13.6]) |

Abbreviations: BMI, body mass index; CI, confidence interval; MI, myocardial infarction; US, United States
